# Thromboembolism risk among patients with diabetes/stress hyperglycemia and COVID-19

**DOI:** 10.1101/2021.04.17.21255540

**Authors:** Stefania L Calvisi, Giuseppe A Ramirez, Marina Scavini, Valentina Da Prat, Giuseppe Di Lucca, Andrea Laurenzi, Gabriele Gallina, Ludovica Cavallo, Giorgia Borio, Federica Farolfi, Maria Pascali, Jacopo Castellani, Vito Lampasona, Armando D’Angelo, Giovanni Landoni, Fabio Ciceri, Patrizia Rovere Querini, Moreno Tresoldi, Lorenzo Piemonti

## Abstract

**Purpose:** Individuals with diabetes/stress hyperglycemia carry an increased risk for adverse clinical outcome in case of SARS-CoV-2 infection. The purpose of this study was to evaluate whether this risk is, at least in part, modulated by an increase of thromboembolic complications.

**Methods:** We prospectively followed 180 hospitalized patients with confirmed COVID-19 pneumonia admitted to the Internal Medicine Units of San Raffaele Hospital. Data from 11 out of 180 patients were considered incomplete and excluded from the analysis. We analysed inflammation, tissue damage biomarkers, hemostatic parameters, thrombotic events (TEs) and clinical outcome according to the presence of diabetes/stress hyperglycemia.

**Results:** Among 169 patients, 51 (30.2%) had diabetes/stress hyperglycemia. Diabetes/stress hyperglycemia and fasting blood glucose (FBG) were associated with increased inflammation and tissue damage circulating markers, higher D-dimer levels, increased prothrombin time and lower antithrombin III activity. Forty-eight venous and 10 arterial TEs were identified in 49 (29%) patients. Diabetes/stress hyperglycemia (HR 2.71, p=0.001), fasting blood glucose (HR 4.32, p<0.001) and glucose variability (HR 1.6, p < 0.009) were all associated with an increased risk of thromboembolic complication. TEs significantly increased the risk for an adverse clinical outcome only in the presence of diabetes/stress hyperglycemia (HR 3.05, p=0.01) or fasting blood glucose ≥ 7 mmol/l (HR 3.07, p=0.015).

**Conclusions:** Thromboembolism risk is higher among patients with diabetes/stress hyperglycemia and COVID-19 pneumonia and is associated to poor clinical outcome. In case of SARS-Cov-2 infection patients with diabetes/stress hyperglycemia could be considered for a more intensive prophylactic anticoagulation regimen.

## Introduction

Diabetes has been confirmed as one of the most consistent risk factors for severe disease in case of SARS-CoV-2 infection^1, 2^. In fact, the risk of admission to an Intensive Care Unit (ICU) and of in-hospital mortality are increased two to three fold by the presence of diabetes in patients with COVID 19 pneumonia^3^. Different pathophysiological mechanisms were suggested to explain the worse clinical outcome, including hyperglycemia, older age and the presence of comorbidities (i.e., hypertension, obesity, and cardiovascular disease)^4^. However, because of the syndromic nature of diabetes, additional potential causative factors should be considered, such as the increased susceptibility to hyperinflammation^5^, the diminished immunological function ^6^ and the prothrombotic state^7^ associated with hyperglycemia. We recently investigated whether diabetes or hyperglycemia are linked to a defect in the humoral immune response against SARS-CoV-2^8, 9^. Our data showed that the antibody response against multiple SARS-CoV-2 antigens in patients with diabetes is superimposable in terms of timing, persistence, classes, titers, and neutralizing activity to that of non-diabetic patients^9, 10^. However, in our cohort of patients with SARS-CoV-2 pneumonia, we also observed a significant correlation between serum D-dimer levels and diabetes/hyperglycaemia, a finding confirmed also by others^11-13^. Elevated D-dimer levels are a direct consequence of increased fibrin formation and lysis and thus an indicator of increased thrombotic activity, such as disseminated intravascular coagulation (DIC) and thromboembolism^14^. COVID-19 is associated with an increased risk of arterial and venous thrombosis ^15-17^ because of a multitude of factors, including systemic inflammation, endothelial dysfunction, platelet activation, immobilization, mechanical ventilation and the use of central venous catheters^18-20^. Since diabetes is associated with a pro-thrombotic status ^7^ and elevated D-dimer levels ^12^, we hypothesized that diabetes is associated with an increased risk of thrombotic events (TEs) in patients with COVID-19 pneumonia. To prove this hypothesis we designed a prospective observational study in a cohort of 180 consecutively hospitalized patients with COVID-19 pneumonia, focusing on TEs which occurred during hospitalization and risk factors associated with these events.

## Material and methods

### Study population and data sources

The study population consisted of 180 adult patients (≥18 years) with confirmed COVID-19 pneumonia admitted to the Internal Medicine Units of San Raffaele Hospital, Milan, Italy from April to May 2020. Patients were included if they were diagnosed with COVID-19 as per the Chinese management guidelines and the World Health Organization interim guidance ^21, 22^. There was no exclusion criterion. A confirmed infection case was defined as a SARS-CoV-2-positive RT-PCR test from a nasal/throat swab, and/or signs, symptoms and radiological findings suggestive of COVID-19 pneumonia. Within 48 hours from admission, we recorded demographic information, clinical features and laboratory exams on the day of admission on a dedicated data collection form. Data were recorded until hospital discharge or death, whichever occurred first. Data were cross-checked in blind and verified by data managers and clinicians for accuracy. We also recorded mortality beyond hospital discharge clinic: for patients non attending our dedicated outpatient follow-up clinic, we checked patient’s vital status with either family members or family physician. The study was approved by the local Institutional Review Board (protocol n 34/int/2020; NCT04318366). A standard written informed consent was requested to all patients for their data use.

### Thrombotic complications

The occurrence of any thrombotic event (TE) throughout the hospitalization was the primary outcome of the study. Thrombotic complications included deep vein thrombosis (DVT), pulmonary embolism (PE), and lower and upper limb ischemia, catheter-related thrombosis with deep vein involvement, mesenteric ischemia, stroke and myocardial infarction. A standard protocol to assess patients for thrombotic complications was implemented based on the position paper from the Italian Society on Thrombosis and Haemostasis (SISET) ^23^ and to the interim guidance to recognition and management of coagulopathy in COVID-19 from the International Society on Thrombosis and Hemostasis (ISTH) ^24^. A close control of hemostasis parameters and clinical signs and symptoms was methodically pursued. Additional investigations, including CT scan and/or ultrasound, were performed on the basis of clinical suspicion of thromboembolic events: (i) elevated D-dimer levels and/or (ii) presence of respiratory failure and/or (iii) presence of symptoms suggestive of TEs. All patients received thromboprophylaxis with enoxaparin 4,000 IU/day [adjusted to 6,000 IU/day or 3,000 IU/day in overweight (>100 kg) or underweight (<50kg) subjects, respectively] or, alternatively, with mechanical compression of the lower limbs in case of anticoagulant contraindications (active bleeding and platelet count less than 25 × 109/l). If chronic oral anticoagulant therapy with direct oral anticoagulants (DOACs) or warfarin/acenocoumarol (OAT) was prescribed prior to admission, it was changed to LMWH anticoagulant treatment. Thromboprophylaxis was administered on admission and during the entire duration of the hospital stay. Anti-Xa measurement was used to monitor anticoagulant treatment. There were no cases of heparin-induced thrombocytopenia. No major haemorrhagic event occurred in patients with thromboprophylaxis. The Padua Prediction Score and the IMPROVE Bleeding Risk Assessment Score were used at hospital admission for stratification of the venous thromboembolism and bleeding risks, respectively. A Padua score ≥ 4 identified patients at high risk for venous thromboembolism, an IMPROVE Bleeding Risk Assessment Score ≥ 7 identified patients at increased risk of bleeding. Overt DIC was defined when the ISTH diagnostic score was ≥ 5 ^25^.

### Definition of diabetes/stress hyperglycemia

Study participants were defined as having diabetes/stress hyperglycaemia if they had a documented diagnosis before the hospital admission for COVID-19 pneumonia [Comorbid diabetes: fasting plasma glucose (FPG) ≥7.0 mmol/l or HbA_1c_ ≥ 6.5% (48 mmol/mol), or prescription for diabetes medications] or if patients without a previous diagnosis of diabetes had a mean FPG ≥7.0 mmol/l. during the hospitalization for COVID-19 pneumonia (stress hyperglycaemia). We computed mean FPG and glucose variability (standard deviation) from all fasting laboratory glucose values measured during hospitalisation.

### Laboratory variables

Routine blood tests encompassed serum biochemistry [including renal and liver function, lactate dehydrogenase (LDH) and electrolytes], complete blood count with differential, markers of myocardial damage [troponin T and pro-brain natriuretic peptide (proBNP)], inflammation markers [C-reactive protein (CRP), ferritin, interleukin-6 (IL-6)] and coagulation profile assessment (D-dimer, PT, and PTT). Specific antibodies to different SARS-CoV-2 antigens were tested in a subset of patients by a luciferase immunoprecipitation system (LIPS) assay, as previously described (11). Fibrinogen, antithrombin activity, vWF, homocysteine, protein C and S. D-dimer levels were measured in a subset of patients through a STA-R® automatic coagulation analyser. Age-specific high D-dimer (aD-dimer) was defined as D-dimer levels above 0.5 µg/dl for patients with less than 50 years of age and above their age divided by 100 in patients older than 50 years^26^.

### Statistical analysis

Continuous variables were presented as median with inter-quartile range (IQR) in parenthesis. Categorical variables were reported as frequency or percent. Continuous variables were compared using the Wilcoxon rank sum or Kruskal-Wallis test. Categorical variables were compared using the Chi-square or Fischer’s exact test, as appropriate. Imputation for missing data was not performed. Associations between baseline variables and diabetes was assessed by logistic regression. The effect estimates were reported as Odd Ratios (ORs). Survival was estimated according to Kaplan–Meier. The time-to-event was calculated from the date of symptom onset to the date of the event, or of last follow-up visit, whichever occurred first. We calculated univariate and multivariate Cox proportional hazards models to study the association between patient characteristics with time to thrombotic complication or time to adverse outcome (as defined by composite endpoint of transfer to ICU or death, whichever occurred first). In Cox proportional hazards models, the onset of a thrombotic complication was considered a time-varying covariate. The effect estimates were reported as Hazard Ratios (HRs) with the corresponding 95% CI, estimated according to the Wald approximation. Multivariate analyses were performed including variables significant at the level of <0.1 in the univariate analysis. Two-tailed P values are reported, with P value <0.05 indicating statistical significance. All confidence intervals are two-sided and not adjusted for multiple testing. Statistical analyses were performed with the SPSS 24 (SPSS Inc. /IBM) and the R software version 3.4.0 (R Core Team (2020).

## Results

### Study participants

A total of 180 consecutive patients with confirmed COVID-19 were prospectively enrolled. Data from 11 out of 180 (6.11%) patients were considered incomplete and excluded from the analysis. Among the 169 cases included in our study [median hospital stay 17 (8-31) days], 61 patients (36.1%) were treated with non-invasive ventilation and 23 (13.6%) accessed an ICU over the hospitalization period. As of January 25, 2021 the median follow-up time after symptoms onset was 222 (95% CI: 211-232) days. Thirty five patients died during follow-up (20.7%). Fifty patients (29.6%) had an adverse in-hospital outcome, according to the composite endpoint of transfer to ICU or death, whichever occurred first.

### Baseline characteristics of study population

The characteristics of study participants, according to diabetes status or glucose levels, are reported in Supplementary Table 1 and Table 2. Stress hyperglycaemia and comorbid diabetes accounted for 11.2% (*n*=19) and 18.9% (*n*=32) of the patients, respectively. Higher BMI [OR 1.112 (95% IC 1.03-1.2); p=0.007], older age [OR 1.029 x year (95% IC 1.01-1.05); p=0.013], and hypertension [OR 4.036 (1.04-3.98); p=0.037] were all associated with diabetes/stress hyperglycaemia. As for diabetes treatment, 3.9% of subject with comorbid diabetes were being treated with lifestyle modifications, 11.8% with insulin, 39.2% with non-insulin oral or injectable anti-diabetes medications, 7.8% with insulin and oral diabetes medications, while patients with stress hyperglycaemia (37.3%) were untreated. The median time from symptoms onset to hospital admission was 7 (1-12.5) and 5 (1-8) days for patients without and with diabetes/stress hyperglycaemia, respectively (p=0.33). On admission, 18.3% (n=31) of the patients were taking ACE-inhibitors (25.5% vs 15.3%, p=0.132 diabetes/stress hyperglycaemia vs no diabetes), 14.2% (n=24) chronic antiplatelet therapy (21.6% vs 11%, p=0.092 diabetes/stress hyperglycaemia vs no diabetes) and 18.3% (n=31) anticoagulant treatments (23.5% vs 16.1%, p= 0.28 diabetes/stress hyperglycaemia vs no diabetes).

### Hospital admission

On admission signs of respiratory insufficiency were evident in most patients [PaO2/FiO2 ratio 280 (200-368)] and a PaO2/FiO2 ratio <200 was present in 20% and 29.4% of patients with or without diabetes/stress hyperglycaemia, respectively (p=0.31). Diabetes/stress hyperglycaemia was associated with worse kidney function [serum creatinine: 96.4 (65.4-152) vs 82.2 (65.4-106) μmol/L, p=0.039; urea nitrogen 18.6 (10.5-33.8) vs 11.9 (8.66-19.6) mmol/L, p=0.004], increased inflammation [CRP 87.5 (35.7-184) vs 53.5 (17.9-112) mg/L; p= 0.009)] and tissue damage markers [LDH 6.65 (4.33–8.8) vs 5.04 (3.82–8.8) μkat/L, p=0.006; AST 0.82 (0.64–1.35) vs 0.6 (0.43–0.91) μkat/L, *p*=0.006; ALT 0.73 (0.45– 0.97) vs 0.57 (0.33–0.83) μkat/L, *p*=0.077; pro-BNP 738 (193-2238) vs 193 (59-910) ng/L, p= 0.011; troponin T 19.5 (11.4-61.55) vs 12.7 (6-42.6) μg/L, p= 0.078]. The same changes were associated with progressively higher blood glucose levels (Supplementary Table 2). Data regarding the IgG, IgM and IgA responses to the SARS-CoV-2 spike protein (RBD or S1+S2) and IgG to NP (Table 1) were available for a subgroup of patients, as they were part of a previous cohort evaluated for the humoral response in the presence of diabetes (12). Marginal differences between patients with and without diabetes/stress hyperglycaemia were evident. Over the hospitalization period antibiotic (80.4% vs 61.9%, p=0.02) and oxygen (84.3% vs 61%, p=0.004) treatments were more frequently used in patients with diabetes/stress hyperglycaemia while antiviral, immunomodulatory and biologic therapies were equally prescribed.

**Table 1.** Hemostatic parameters, inflammation markers and SARS-Cov2 antibodies according to diabetes/stress hyperglycemia and median fasting plasma glucose.

| Characteristics | All | Diabetes/stress hyperglycemia |  |  | Median fasting glucose (mmol/l) |  |  |  | Missing |
| --- | --- | --- | --- | --- | --- | --- | --- | --- | --- |
|  |  | No | Yes | p | <5.6 | 5.6-6.9 | ≥ 7 | p |  |
| N |  | 118 | 51 |  | 90 | 37 | 42 |  |  |
| Platelets (x10<sup>9</sup>/L) | 225 (153-307) | 230 (157-309) | 212 (151-301) | 0.61 | 229 (157-379) | 200 (160-321) | 225 (146-324) | 0.90 | 1 |
| D-dimer (µg/ml) | 5.9 (2.8-16.6) | 4.2 (2.3-9.3) | 11.8 (5.5-12.4) | <0.001 | 4.16 (2.3-9) | 5.75 (2.35-11.4) | 15.1 (6-35.6) | <0.001 | 0 |
| Elevated aD-dimer [N (%)] | 106 (62.7) | 67 (56.8) | 39 (76.5) | 0.016 | 51 (56.7) | 21 (56.8) | 34 (81) | 0.019 | 0 |
| C reactive protein (mg/l) | 57.5 (20.7-128.6) | 53.5 (17.9-112) | 87.5 (35.7-184) | 0.009 | 35.4 (11.7-92.5) | 69.1 (54-130) | 117 (47.2-204) | <0.001 | 4 |
| PT-INR | 1.06 (0.99-1.2) | 1.04 (0.98-1.15) | 1.16 (1.02-1.25) | 0.001 | 1.04 (0.98-1.17) | 1.06 (0.97-1.16) | 1.17 (1.04-1.27) | 0.005 | 4 |
| PTT-R | 0.99 (0.94-1.05) | 1 (0.95—1.05) | 0.98 (0.91-1.06) | 0.52 | 0.99 (0.94-1.05) | 1.01 (0.95-1.1) | 0.97 (0.9-1.06) | 0.27 | 4 |
| Ferritin µg/L | 903 (387-1514) | 804 (331-1450) | 1058 (484-1689) | 0.17 | 700 (308-1115) | 1367 (447-2492) | 1092 (588-1821) | 0.003 | 29 |
| IL-6 (pg/ml) | 43.3 (15.4-98.5) | 43.7 (13.2-95.85) | 41.5 (22-124) | 0.26 | 39.2 (9.5-86) | 74.3 (13.4-131) | 41.4 (26.7-153) | 0.12 | 44 |
| Fibrinogen (g/L) | 5.53 (4.19-6.39) | 5.46 (4.16-6.19) | 5.65 (4.18-7.34) | 0.24 | 5.11 (3.89-5.85) | 5.87 (5.57-6.59) | 5.75 (4.78-7.46) | 0.004 | 42 |
| von Willebrand factor (%) | 307 (190-379) | 306 (212-377) | 336 (165-420) | 0.92 | 307 (191-376) | 301 (176-372) | 358 (184-420) | 0.49 | 101 |
| Protein C (%) | 86 (72-106) | 86 (77-103) | 73.5 (53-111) | 0.24 | 87 (76-104) | 82.5 (72-93) | 78.5 (53-111) | 0.59 | 96 |
| Protein S (%) | 78 (60-98) | 82 (60-98) | 68 (56-94) | 0.36 | 84 (61.5-101.5) | 78.5 (59-94.5) | 68 (63-88.5) | 0.56 | 98 |
| Antithrombin III (%) | 98 (88-106) | 100 (92-106) | 91 (80-102) | 0.041 | 100 (88.25-105) | 98 (93-109) | 89 (79-104) | 0.23 | 95 |
| Homocysteine (µmol/L) | 14.8 (10.1-19.4) | 14.4 (10-18.4) | 16.5 (12.3-23.5) | 0.17 | 13.4 (10-16) | 19.4 (10-24) | 15.4 (9.9-21.7) | 0.140 | 102 |
| Padua score | 3 (2-5) | 3(1-5) | 3 (3-5) | 0.006 | 3 (1-5) | 3 (1.5-4.5) | 3 (3-5) | 0.049 | 0 |
| Padua score ≥ 4 [N (%)] | 59 (34.9) | 37 (31.4) | 22 (43.1) | 0.16 | 30 (33.3) | 11 (29.7) | 18 (42.9) | 0.43 | 0 |
| IMPROVE score | 2 (1-3) | 1.6 (0.97-2.9) | 2.2 (1.2-4.1) | 0.024 | 1.6 (0.9-2.9) | 1.5 (11-2.9) | 2.2 (1.42-4.02) | 0.15 | 0 |
| IMPROVE score ≥ 7 [N (%)] | 12 (7.1) | 7 (5.9) | 5 (9.8) | 0.35 | 7 (7.8) | 1 (2.7) | 4 (9.5) | 0.47 | 0 |
| DIC by ISTH definitions [N (%)] | 4 (2.4) | 1 (0.8) | 3 (5.9) | 0.083 | 0 (0) | 1 (2.7) | 3 (7.1) | 0.042 | 0 |
| IgG RBD [N (%)] | 45 (51.1) | 30 (47.6) | 15 (60) | 0.349 | 24 (50) | 8 (47.1) | 13 (56.5) | 0.817 | 81 |
| IgM RBD [N (%)] | 46 (52.3) | 33 (52.4) | 13 (52) | 0.99 | 24 (50) | 10 (58.8) | 12 (52.2) | 0.822 | 81 |
| IgA RBD [N (%)] | 44 (50) | 27 (42.9) | 17 (68) | 0.057 | 19 (39.6) | 9 (52.9) | 16 (69.6) | 0.059 | 81 |
| IgG S1/S2 | 53 (60.2) | 36 (57.1) | 17 (68) | 0.470 | 28 (58.3) | 10 (58.8) | 15 (65.2) | 0.850 | 81 |
| IgM S1/S2 | 61 (69.3) | 42 (66.7) | 19 (76) | 0.452 | 33 (68.8) | 11 (64.7) | 17 (73.9) | 0.816 | 81 |
| IgA S1/S2 | 66 (75) | 45 (71.4) | 21 (84) | 0.281 | 34 (70.8) | 13 (76.5) | 19 (82.6) | 0.556 | 81 |
| IgG NP | 58 (65.9) | 42 (66.7) | 16 (64) | 0.808 | 31 (64.6) | 12 (70.6) | 15 (65.2) | 0.901 | 81 |

DIC disseminated intravascular coagulation; INR: international normalized ratio ISTH: International Society of Thrombosis and Haemostasis; PT prothrombin time; PTT: partial thromboplastin time.

### Hemostatic parameters

Upon admission patients with diabetes/stress hyperglycaemia exhibited significantly higher D-dimer levels [11.8 (5.5-12.4) vs 4.2 (2.3-9.3) µg/ml, p<0.001] and increased prothrombin time (PT-INR 1.16 (1.02-1.25 vs 1.04 (0.98-1.15), p=0.001) compared with patients without diabetes/stress hyperglycaemia (Table 1). Concordantly, the percentage of subject with elevated age-specific D-dimer was significantly higher in the presence of diabetes/stress hyperglycaemia (76.5% vs 56.8%, p=0.016). Partial thromboplastin time, platelet count and fibrinogen were not affected by diabetes/stress hyperglycaemia. Advanced markers of thrombophilia were available for a subgroup of patients (Table 1). Exploratory analysis did not show significant changes in coagulation factors levels or activity in subjects with diabetes, except for a lower antithrombin III activity [91% (80-120) vs 100% (92-106)]. The presence of diabetes/stress hyperglycaemia did not significantly increase the proportion of patients who at baseline had a higher risk for venous thromboembolism based on the Padua score (Padua score ≥ 4: 43.1% vs 31.4%, p=0.16), even if a marginal, although significant, difference in the Padua score was evident [3 (3-5) vs 3 (1-5), p=0.006)] (Table 1). Similarly, the proportion of patients at high baseline bleeding risk based on the IMPROVE score was not affected by the presence of diabetes/stress hyperglycaemia (IMPROVE≥ 7: 9.8% vs 5.9%; p= 0.35), even if a marginal, although significant, difference in the IMPROVE score was evident [2.2 (1.2-4.1) vs 1.6 (0.97-2.9), p=0.024)] (Table 1).

### Thromboembolic complications

Forty eight venous and 10 arterial TE were identified in 49 (29%) patients (Table 2). The median time from the onset of symptoms of COVID-19 pneumonia to the TE was 17 (10-24) days. Patients with diabetes/stress hyperglycaemia developed more frequently a thromboembolic complication (47.1% vs 21.2%, p=0.001) than patients without diabetes/stress hyperglycaemia. Comorbid diabetes and stress hyperglycaemia were both associated with a higher prevalence of thromboembolic complication (52% vs 42.1% respectively, p=0.77). Concordantly, the higher prevalence of thromboembolic complication was associated with higher fasting blood glucose levels during the hospitalization (Table 1). Regarding the thrombosis site, venous events (in particular deep vein thrombosis) contributed more significantly than arterial ones in determining the higher prevalence of thromboembolic complications in patients with diabetes/stress hyperglycaemia compared to those without diabetes/stress hyperglycaemia. In patients with and without diabetes/stress hyperglycaemia, 3 and 1 (5.9% vs 0.8%, p=0.083) events were classified as overt DIC (≥5 points, according to the ISTH diagnostic criteria), respectively. The results of a Cox regression analysis for TE is presented in Fig. 1. The Cox regression analysis adjusted for age and sex indicates that diabetes/stress hyperglycaemia (HR 2.71, CI 1.53-4.8; p=0.001), fasting plasma glucose [FPG mean (log1p) HR 4.32, CI 1.86-10, p=0.001] and glucose variability [FPG standard deviation (log1p) HR 1.6, CI 1.13-2.28, p=0.009] were associated with a higher risk of thromboembolic complications. Differences in smoke habit, BMI, other comorbidities and preadmission antiplatelet or steroid were all statistically not significant. A trend towards a protective effect of preadmission anticoagulant therapy was evident (HR 0.45, CI 0.18-1.08; p=0.075). In a multivariate model, diabetes/stress hyperglycaemia and preadmission anticoagulant therapy confirmed their association with the thromboembolic complication.

**Table 2:** Thrombotic events (TEs)

|  | Diabetes/stress hyperglycemia |  |  |  |  |  | Median fasting glucose (mmol/l) |  |  |  |  |
| --- | --- | --- | --- | --- | --- | --- | --- | --- | --- | --- | --- |
|  | All | No | Yes | p | Comorbid diabetes | Stress hyperglycemia | p | <5.6 | 5.6-6.9 | ≥ 7 | p |
| **N** | 169 | 118 | 51 |  | 32 | 19 |  | 90 | 37 | 42 |  |
| **Patients with at least one event [N (%)]** | 49 (29) | 25 (21.2) | 24 (47.1) | 0.001 | 16 (50) | 8 (42.1) | 0.77 | 20 (22.2) | 9 (24.3) | 20 (47.6) | 0.009 |
| Median time from symptoms to thrombosis | 17 (10-24) | 11 (2-20) | 18 (8-28) | 0.11 | 21 (11-31) | 11 (0-28) | 0.44 | 13 (4-22) | 6 (0-12) | 18 (3-33) | 0.19 |
| **Arterial events [N (%)]** |  |  |  |  |  |  |  |  |  |  |  |
| - Myocardial infarction | 3 (1.8) | 1 (0.8) | 2 (3.9) | 0.22 | 2 (6.3) | 0 (0) | 0.52 | 0 (0) | 2 (5.4) | 1 (2.4) | 0.10 |
| - Stroke | 1 (0.6) | 0 (0) | 1 (2) | 0.30 | 1 (3.1) | 0 (0) | 0.99 | 0 (0) | 0 (0) | 1 (2.4) | 0.19 |
| - Limb ischaemia | 6 (3.6) | 4 (3.4) | 2 (3.9) | 0.99 | 1 (3.1) | 1 (5.3) | 0.99 | 4 (4.4) | 0 (0) | 2 (4.8) | 0.42 |
| - Intestinal ischaemia | 0 (0) | 0 (0) | 0 (0) | - | 0 (0) | 0 (0) | - | 0 (0) | 0 (0) | 0 (0) | - |
| Total patients with arterial event | 10 (5.9) | 5 (4.2) | 5 (9.8) | 0.17 | 4 (12.5) | 1 (5.3) | 0.64 | 4 (4.4) | 2 (5.4) | 4 (9.5) | 0.51 |
| **Venous events [N (%)]** |  |  |  |  |  |  |  |  |  |  |  |
| - Isolated spontaneous DVT | 17 (10.1) | 6 (5.1) | 11 (21.6) | 0.004 | 8 (25) | 3 (15.8) | 0.50 | 6 (6.7) | 3 (8.1) | 8 (19) | 0.08 |
| - Isolated PVT | 15 (8.9) | 8 (6.8) | 7 (13.7) | 0.15 | 4 (12.5) | 3 (15.8) | 0.99 | 4 (4.4) | 5 (13.5) | 6 (14.3) | 0.096 |
| - DVT+ PVT | 8 (4.7) | 6 (5.1) | 2 (3.9) | 0.99 | 1 (3.1) | 1 (5.3) | 0.99 | 6 (6.7) | 0 (0) | 2 (4.8) | 0.27 |
| - Catheter-related DVT | 8 (4.7) | 3 (2.5) | 5 (9.8) | 0.055 | 4 (12.5) | 1 (5.3) | 0.64 | 4 (4.4) | 1 (2.7) | 3 (7.1) | 0.64 |
| Total patients with venous events | 43 (25.4) | 22 (18.6) | 21 (48.8) | 0.004 | 13 (40.6) | 8 (42.1) | 0.99 | 18 (20) | 8 (21.6) | 17 (40.5) | 0.035 |
| **Arterial and venous events [N (%)]** |  |  |  |  |  |  |  |  |  |  |  |
| - Myocardial infarction + DVT/PVT | 1 (0.6) | 0 (0) | 1 (2) | 0.3 | 1 (3.1) | 0 (0) | 0.99 | 0 (0) | 1 (2.7) | 0 (0) | 0.17 |
| - Stroke + DVT/PVT | 0 (0) | 0 (0) | 0 (0) | - | 0 (0) | 0 (0) | - | 0 (0) | 0 (0) | 0 (0) | - |
| - Limb ischemia + DVT/PVT | 3 (1.8) | 2 (1.7) | 1 (2) | 0.99 | 0 (0) | 1 (5.3) | 0.37 | 2 (2.2) | 0 (0) | 1 (2.4) | 0.65 |
| Total patients with arterial and venous events | 4 (2.4) | 2 (1.7) | 2 (3.9) | 0.58 | 1 (3.1) | 1 (5.3) | 0.99 | 2 (2.2) | 1 (2.7) | 1 (2.4) | 0.99 |

DVT: Deep vein thrombosis; PVT: Pulmonary vein thrombosis

**Figure 1.**
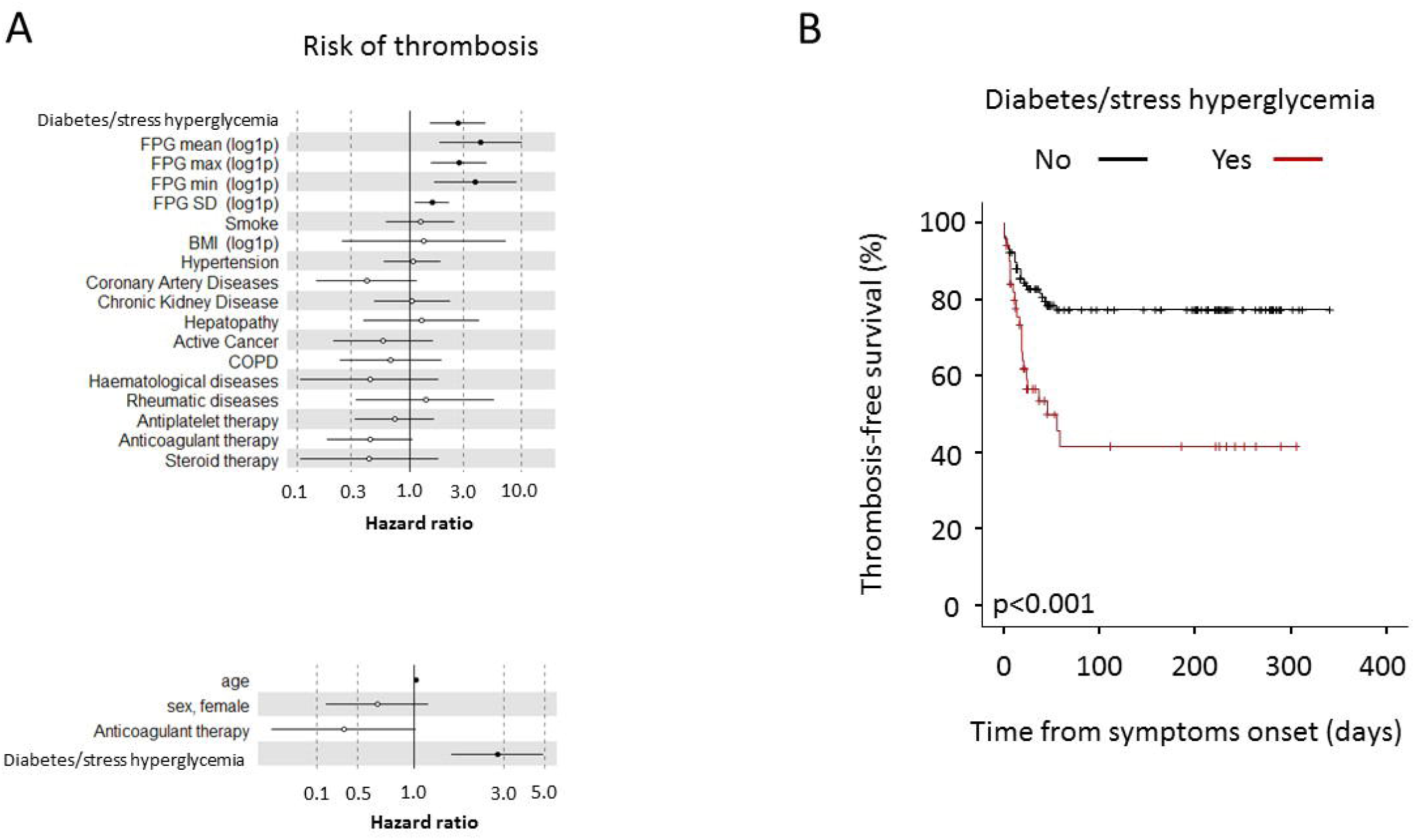
TEs in patients with COVID-19. The forest plots (panel a) show the Hazard Ratios (HR) for thrombosis for each factor tested. Upper panel: univariate Cox regression analysis adjusted for sex and age. Lower panel: multivariate Cox regression analysis adjusted for sex and age including variables significant at the level of <0.1 in the univariate analysis. Dots represent the HR, lines represent 95% confidence interval (CI), and solid dots indicate P < 0.05. Kaplan-Meier thrombosis-free survival estimates for patients with COVID-19 pneumonia (panel b). Survival rate was estimated for the presence of diabetes. The log-rank test was used to test differences in the estimated survival rate. Crosses indicate censored patients (censoring for death end of follow-up data).

### TEs and adverse clinical outcome in patients with and without diabetes/stress hyperglycaemia

To assess whether the presence of TEs had an impact on patient outcome according to diabetes/stress hyperglycaemia or fasting glucose levels, we conducted a Kaplan**-** Meier estimator log-rank test and a Cox proportional hazards model for adverse clinical outcome (as defined by composite endpoint of transfer to ICU or death, whichever occurred first) (Figure 2). The Cox regression analysis adjusted for age and ***sex*** indicated that diabetes/stress hyperglycaemia (HR 2.99, CI 1.7-5.03; p<0.001), fasting plasma glucose [FPG mean (log1p) HR 9.6, CI 4.59-20; p<0.001] and glucose variability [FPG standard deviation (log1p) HR 2.02, CI 1.43-2.9; p<0.001] were strongly associated with a higher risk of adverse clinical outcome (see also Supplementary Table 2). TEs were not associated with an adverse clinical outcome in the absence of diabetes/stress hyperglycaemia (HR 0.29, CI 0.04-2.16; p=0.225) or in the presence of FBG <7 mmol/l (HR 0.54, CI 0.07-4.35; p=0.56), while they significantly increased the risk in the presence of diabetes/stress hyperglycaemia (HR 3.05, CI 1.31-7.09; p=0.01) or FBG ≥ 7 mmol/l (HR 3.07, CI1.24-7.6; p=0.015).

**Figure 2.**
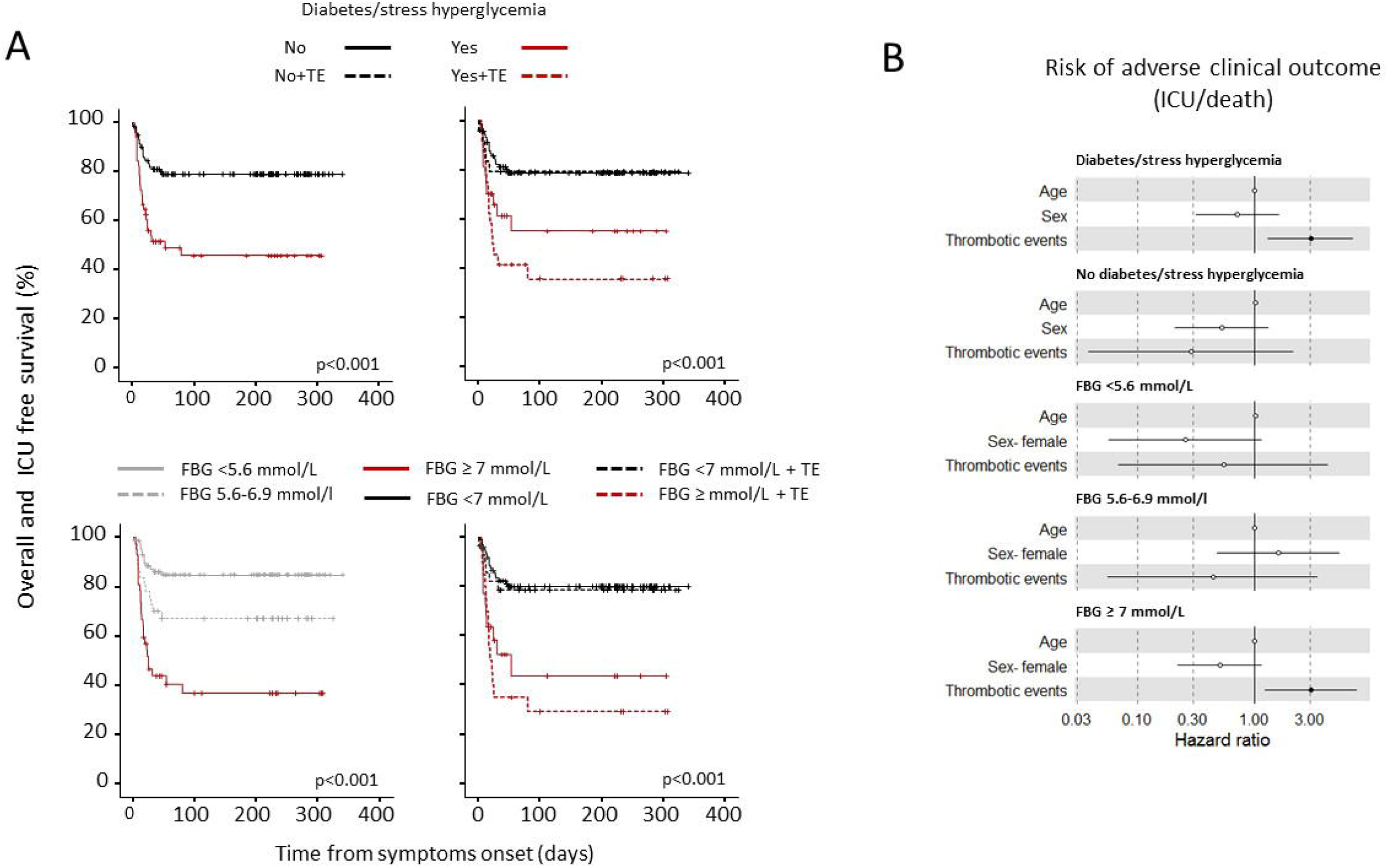
Survival in the absence of adverse clinical outcome in patients with COVID-19 with or without TEs, according to diabetes/stress hyperglycemia or fasting glucose levels. Kaplan-Meier patient survival estimates for 169 patients with COVID-19 pneumonia (panel a). Survival rate in the absence of adverse clinical outcome (defined by composite endpoint of transfer to ICU or death, whichever occurred first) was estimated for the presence of any thrombotic event (TE) separately according diabetes/stress hyperglycemia or fasting glucose levels. The log-rank test was used to test differences in the estimated survival rate between groups. Crosses indicate censored patients (censoring for death or end of follow-up data). The forest plots (panel b) show the hazard ratios for survival in the absence of adverse clinical outcome according to presence/absence of diabetes/stress hyperglycemia or fasting glucose levels (FBG) categories. The presence of thrombotic events (TE) was considered as a time-varying covariate in Cox proportional hazards models. The effect estimates were reported as Hazard Ratios (HRs) with the corresponding 95% CI, estimated according to the Wald approximation. Cox regression analysis was adjusted for sex and age. Dots represent the HR, lines represent 95% confidence interval (CI), and solid dots indicate P < 0.05.

## Discussion

There are few reports on the relationship between hyperglycemia and the rate of TEs in COVID-19 pneumonia and it is still unknown whether thrombosis affects the prognosis of patients with COVID-19 pneumonia in the presence of diabetes. Under the hypothesis of a relevant role for diabetes, we designed a prospective observational study focusing on TEs occurring during hospitalization and risk factors associated with thromboembolic complications in patients with COVID-19 pneumonia. Our study generated several interesting findings in those patients. First, diabetes/stress hyperglycaemia, high fasting glycaemia and glycemic variability were strong risk factors for the development of thromboembolic complications. Second, the rate of venous thrombosis events (in particular deep vein thrombosis) was the most affected by the presence of diabetes/stress hyperglycaemia. Third, thromboembolic complications had an adverse impact on clinical outcome exclusively in the presence of diabetes/stress hyperglycaemia. While reasonable, these results could not have been taken for granted^27^. TEs have a higher incidence among patients with COVID-19 ^28-30^ and diabetes is *per se* characterized by a pro-thrombotic status ^7^. We and others have previously reported an increase of D-dimer in patients with diabetes/stress hyperglycaemia and COVID-19 pneumonia compared to those without ^11-13^. However the clinical implications in term of thromboembolic risk of those findings were yet unclear. Moreover, data on the correlation of thromboembolic complications with clinical outcome were limited and contradictory, with some studies finding a higher risk of adverse outcome associated with TEs in hospitalized patients with COVID-19 ^31^, while others did not find any association ^32^. The pathophysiological mechanisms related to the increased risk of TEs in patients with COVID-19 pneumonia and diabetes are still incompletely understood. In our study, diabetes/stress hyperglycaemia was associated with both inflammation and coagulopathy (elevated C reactive protein and D-dimer levels, mild prolongation of the prothrombin time and decreased antithrombin III), suggesting that an hyperglycaemia-related amplification of the pathobiological mechanisms of immunothrombosis ^33^ could be responsible of the increased thrombotic risk. The reduced activity of antithrombin III is of particular interest in this context ^34^. In fact, antithrombin III is a powerful natural anticoagulant which is regulated by inflammation ^35^. Therefore, it can be speculated that hyperinflammation might have been triggering a decrease in antithrombin III levels and its physiological anticoagulant activity. Furthermore, since the clinical anticoagulant efficacy of heparin requires interaction with antithrombin III, an impaired levels/activity of antithrombin III may be associated with “heparin resistance” ^35^. In agreement with this hypothesis, an association between antithrombin III levels and mortality in patients with COVID-19 pneumonia has already been reported^36^. An association between thromboembolic complications and the presence of antiphospholipid antibodies in patients with COVID-19 pneumonia has been previously reported in case reports, case series, cohort studies, and cross-sectional studies, although with contradictory results ^37^. Unfortunately antiphospholipid antibody measurements were available only for a subgroup of our patients and, therefore, it was impossible to include them in a multivariate model to test for their contribution to thromboembolic risk.

Our study encompasses some limitations: first, our cohort was limited to hospitalized patients and results could be different in less severe COVID-19 disease. Second, even if the overall venous and arterial thromboembolism rate was similar to that described until now in various studies (18), our monocentric cohort was relatively small, and, therefore, a selection bias cannot be excluded. Third, we were unable to evaluate the specific role of some markers as predictors of thrombosis in multivariate models since a complete set of biochemical coagulation data was available only for a fraction of patients. Nevertheless our study generated additional valuable knowledge about the role of diabetes/stress hyperglycaemia in predicting TEs and in stratifying their prognostic significance. In conclusion, many evidences indicate that patients with diabetes, in case of COVID-19 pneumonia, carry a significant increased risk for adverse clinical outcome when compared with patients without diabetes. It is clear from our study that part of this risk is due to an increase in thromboembolic complications. These findings suggest that in in case of SARS-Cov-2 pneumonia, patients with diabetes/stress hyperglycaemia could be considered for a more intensive prophylactic anticoagulation regimen.

## Data Availability

Some or all datasets generated during and/or analyzed during the current study are not publicly available but are available from the corresponding author on reasonable request.

https://figshare.com/articles/online_resource/Thromboembolism_risk_is_higher_among_patients_with_diabetes_and_COVID-19_pneumonia_and_is_associated_to_poor_clinical_outcome_an_observational_cohort_study/14381777

## Individual contributions

LP, SLC and GAR contributed to the conception of the study, wrote the manuscript, researched data and contributed to the discussion. MSc and VL contributed to the acquisition and analysis of antibody data and revised the manuscript. VDP, GDL, AL, PRQ and FC recruited patients, contributed to the acquisition of samples, managed the biobanking activities and critically revised the manuscript, AD and GL contributed to the acquisition, analysis and interpretation of data and critically revised the manuscript. MT contributed to the design of the study and critically reviewed/edited the manuscript. LP is the guarantor of this work and, as such, had full access to all the data presented in the study and takes responsibility for the integrity of the data and the accuracy of the data analysis. The final manuscript has been read and approved by all named authors.

