## Supplementary Table 1 for "Thromboembolism risk among patients with diabetes/stress hyperglycemia and COVID-19"

**Supplementary Table 1. Baseline characteristics of study population according to diabetes/stress hyperglycemia and median fasting plasm glucose**

| **Characteristics** | **All** | **diabetes/stress hyperglycemia** | | | **Median fasting glucose (mmol/l)** | | | |  |
| --- | --- | --- | --- | --- | --- | --- | --- | --- | --- |
|  |  | **No** | **Yes** | **p** | **<5.6** | **5.6-6.9** | **≥ 7** | **p** | **Missing** |
| N |  | 118 | 51 |  | 90 | 37 | 42 |  |  |
| Age, years | 66 (53-77.5) | 62 (50-77.5) | 71 (61-78) | 0.014 | 62 (48.7-76.2) | 64 (55-79) | 72 (64-82.5) | 0.021 | 0 |
| Sex Male [N (%)] | 118 (69.8) | 75 (70.8) | 43 (68.3) | 0.73 | 55 (61.1) | 28 (75.7) | 23 (54.8) | 0.14 | 0 |
| Non Caucasian ethnicity [N (%)] | 32 (18.9) | 26 (22) | 6 (11.8) | 0.14 | 21 (23.3) | 5 (13.5) | 6 (14.3) | 0.30 | 0 |
| BMI | 25.9 (23.5-30.2) | 25.6 (23.1-32.3) | 28 (24.7-33.1) | 0.005 | 25.6 (22.8-29.4) | 26.6 (25-29.9) | 27.7 (24.2-31.8) | 0.087 | 56 |
| Smoke [N (%)] | 29 (17.3) | 23 (19.5) | 6 (12) | 0.27 | 19 (21.1) | 7 (18.9) | 3 (7.3) | 0.150 | 1 |
| Previous venous thromboembolism [N (%)] | 9 (5.3) | 6 (5.1) | 3 (5.9) | 0.99 | 3 (3.3) | 3 (8.1) | 3 (7.1) | 0.46 | 0 |
| Previous bleeding [N (%)] | 8 (4.7) | 5 (4.2) | 3 (5.9) | 0.70 | 4 (4.4) | 2 (5.4) | 2 (4.8) | 0.97 | 0 |
| Previous cancer [N (%)] | 13 (7.7) | 9 (7.6) | 4 (7.8) | 0.99 | 9 (10) | 1 (2.7) | 3 (7.1) | 0.37 | 0 |
| Comorbidities [N (%)] |  |  |  |  |  |  |  |  | 0 |
| - Hypertension | 77 (45.6) | 47 (39.8) | 30 (58.8) | 0.029 | 36 (40) | 18 (48.6) | 23 (54.8) | 0.26 |  |
| - Diabetes | 51 (30.2) | 0 (0) | 51 (100) | - | 3 (3.3) | 6 (16.2) | 42 (100) | 0.001 |  |
| - Coronary Artery Diseases | 23 (13.6) | 16 (13.6) | 7 (13.7) | 0.99 | 12 ( 13.3) | 7 (18.9) | 4 (9.5) | 0.47 |  |
| - Active Cancer | 20 (11.8) | 14 (11.9) | 6 (11.8) | 0.99 | 13 (14.4) | 3 (8.1) | 4 (9.5) | 0.52 |  |
| - COPD | 19 (11.2) | 12 (10.2) | 7 (13.7) | 0.60 | 8 (8.9) | 5 (13.5) | 6 (14.3) | 0.59 |  |
| - Chronic Kidney Disease | 18 (10.7) | 9 (7.6) | 9 (17.6) | 0.062 | 7 (7.8) | 3 (8.1) | 8 (19) | 0.13 |  |
| - Haematological diseases | 15 (8.9) | 11(9.3) | 4 (7.8) | 0.99 | 8 (8.9) | 3 (8.1) | 4 (9.5) | 0.98 |  |
| - Hepatopathy | 7 (4.1) | 3 (2.5) | 4 (7.8) | 0.2 | 1 (1.1) | 2 (5.4) | 4 (9.5) | 0.071 |  |
| - Rheumatic disease | 6 (3.6) | 3 (2.5) | 3 (5.9) | 0.37 | 3 (3.3) | 0 (0) | 3 (7.1) | 0.23 |  |
| Preadmission treatment [N (%)] |  |  |  |  |  |  |  |  | 0 |
| - ACE-inhibitor therapy | 31 (18.3) | 18 (15.3) | 13 (25.5) | 0.132 | 16 (17.8) | 6 (16.2) | 9 (21.4) | 0.82 |  |
| - Antiplatelet therapy | 24 (14.2) | 13 (11) | 11 (21.6) | 0.092 | 9 (10) | 7 (18.9) | 8 (19) | 0.25 |  |
| - Anticoagulant therapy | 31 (18.3) | 19 (16.1) | 12 (23.5) | 0.28 | 12 (13.3) | 8 (21.6) | 11 (26.2) | 0.17 |  |
| - DOACs | 10 (5.9) | 6 (5.1) | 4 (7.8) | 0.49 | 6 (6.7) | 1 (2.7) | 3 (7.1) | 0.64 |  |
| - OAT | 5 (3) | 3 (2.5) | 2 (3.9) | 0.64 | 1 (1.1) | 2 (5.4) | 2 (4.8) | 0.31 |  |
| - LMWH | 16 (9.5) | 10 (8.5) | 6 (11.8) | 0.57 | 5 (5.6) | 5 (13.5) | 6 (14.3) | 0.18 |  |
| - Immunosuppression | 4 (2.4) | 4 (3.4) | 0 (0) | 0.32 | 4 (4.4) | 0 (0) | 0 (0) | 0.17 |  |
| - Steroid | 13 (7.7) | 11 (9.3) | 2 (3.9) | 0.35 | 9 (10) | 2 (5.4) | 2 (4.8) | 0.48 |  |
| - Antihyperglycemic agents |  |  |  |  |  |  |  |  |  |
| - Metformin | 20 (11.9) | 0 (0) | 20 (40) | <0.001 | 1 (1.1) | 6 (16.2) | 13 (31.7) | <0.001 |  |
| - Sulfonylureas | 6 (3.6) | 0 (0) | 6 (12) | <0.001 | 1 (1.1) | 0 (0) | 5 (12.2) | 0.003 |  |
| Gliclazide | 4 (2.4) | 0 (0) | 4 (8) | 0.007 | 0 (0) | 0 (0) | 4 (9.8) | 0.002 |  |
| Glibenclamide | 1 (0.6) | 0 (0) | 1 (2) | 0.30 | 0 (0) | 0 (0) | 1 (2.4) | 0.21 |  |
| Glimepiride | 1 (0.6) | 0 (0) | 1 (2) | 0.30 | 1 (1.1) | 0 (0) | 0 (0) | 0.65 |  |
| - Repaglinide | 1 (0.6) | 0 (0) | 1 (2) | 0.30 | 0 (0) | 1 (2.7) | 0 (0) | 0.17 |  |
| - DPP-4 inhibitors | 3 (1.8) | 0 (0) | 3 (6) | 0.025 | 0 (0) | 0 (0) | 3 (7.3) | 0.009 |  |
| - SGLT2 inhibitors | 1 (0.6) | 0 (0) | 1 (2) | 0.30 | 0 (0) | 1 (2.7) | 0 (0) | 0.17 |  |
| - Insulin | 10 (6) | 0 (0) | 10 (20) | <0.001 | 1 (1.1) | 0 (0) | 9 (22) | <0.001 |  |

COPD chronic obstructive pulmonary disease; ACE angiotensin-converting enzyme; DOACs: direct oral anticoagulants;LMWH: Low-molecular-weight heparin; OAT: warfarin/acenocoumarol
