## Supplementary Table 2 for "Thromboembolism risk among patients with diabetes/stress hyperglycemia and COVID-19"

**Supplementary Table 2. Outcomes and laboratory values according to** **diabetes/stress hyperglycemia and median fasting plasma glucose**

| **Characteristics** | **All** | **diabetes/stress hyperglycemia** | | | **Median fasting glucose (mmol/l)** | | | |  |
| --- | --- | --- | --- | --- | --- | --- | --- | --- | --- |
|  |  | **No** | **Yes** | **p** | **<5.6** | **5.6-6.9** | **≥ 7** | **p** | **Missing** |
| N |  | 118 | 51 |  | 90 | 37 | 42 |  |  |
| **Outcomes** |  |  |  |  |  |  |  |  |  |
| Median time from symptoms to admission | 6 (1-11) | 7 (1-12.5) | 5 (1-8) | 0.33 | 7 (0.75-12.2) | 7 (1-12.5) | 5 (1.7-7.5) | 0.77 | 0 |
| Median follow up, days (95%CI) | 222 (211-232) | 220 (209-230) | 231 (214-247) | 0.69 | 213 (192-233) | 231 (217-244) | 231 (215-246) | 0.62 | 0 |
| Hospital stay, days | 17 (8-31) | 14.5 (7-30) | 21 (12-35) | 0.040 | 13.5 (7-27.7) | 21 (8-30) | 22.5 (12.7-35.5) | 0.08 | 0 |
| Non-invasive ventilation | 61 (36.1) | 33 (28) | 28 (54.9) | 0.002 | 18 (20) | 20 (54.1) | 23 (54.8) | <0.001 | 0 |
| Invasive ventilation (ICU) | 23 (13.6) | 11 (9.3) | 12 (23.5) | 0.025 | 5 (5.6) | 7 (18.9) | 11 (26.2) | 0.003 | 0 |
| Death | 35 (20.7) | 15 (12.7) | 20 (39.2) | <0.001 | 9 (10) | 7 (18.9) | 19 (45.2) | <0.001 | 0 |
| Adverse outcome (death and/or Invasive ventilation) | 50 (29.6) | 24 (20.3) | 26 (51) | <0.001 | 13 (14.4) | 12 (32.4) | 25 (59.5) | <0.001 | 0 |
| **Hospital treatment [N (%)]** |  |  |  |  |  |  |  |  |  |
| Antibiotic treatment | 114 (67.5) | 73 (61.9) | 41 (80.4) | 0.02 | 57 (63.3) | 23 (62.2) | 34 (81) | 0.098 | 0 |
| Hydroxychloroquine | 109 (64.5) | 78 (66.1) | 31 (60.8) | 0.6 | 56 (62.2) | 28 (75.7) | 25 (59.5) | 0.26 | 0 |
| Antiviral treatment | 56 (33.1) | 39 (33.1) | 17 (33.3) | 0.99 | 25 (27.8) | 17 (45.9) | 14 (33.3) | 0.14 | 0 |
| Steroids | 32 (19) | 23 (19.5) | 9 (18) | 0.99 | 18 (20) | 5 (13.5) | 9 (22) | 0.60 | 0 |
| Biologics | 27 (16) | 17 (14.4) | 10 (19.6) | 0.49 | 11 (12.2) | 6 (16.2) | 10 (23.8) | 0.25 | 0 |
| Oxigen | 115 (68) | 72 (61) | 43 (84.3) | 0.004 | 49 (54.4) | 29 (78.4) | 37 (88.1) | <0.001 |  |
| **Laboratory parameters** |  |  |  |  |  |  |  |  |  |
| Random fasting glycaemia (mmol/l) |  |  |  |  |  |  |  |  |  |
| - Mean | 97 (85-125) | 89 (81-100) | 147 (132-192) | <0.001 | 86 (80-94) | 111 (104- 116) | 159.5 (139-197) | <0.001 | 0 |
| - Max | 116 (96-173) | 106 (91-122) | 209 (152-265) | <0.001 | 97.5 (87-113) | 128 (114-153) | 227 (187-282) | <0.001 | 0 |
| - Min | 81 (70-97) | 78.5 (68-88) | 108 (82-139) | <0.001 | 75 (675-82) | 90 (81-103) | 120 (88-150) | <0.001 | 0 |
| - SD | 16 (9-30) | 13 (7-18) | 46 (25-57) | <0.001 | 11 (6-16) | 20 (13-29) | 49 (32-72) | <0.001 | 0 |
| - N° of determination | 3 (2-8) | 3 (2-7) | 3 (2-8) | 0.87 | 4 (2-8) | 3 (2-6) | 3 (2-8) | 0.81 | 0 |
| PaO2/FiO2 | 280 (200-368) | 300 (196-395) | 258 (204-349) | 0.25 | 305 (198-400) | 269 (195-363) | 258 (199-349) | 0.32 | 22 |
| - <300 | 80 (54.4) | 52 (51) | 28 (62.2) | 0.22 | 37 (48.7) | 20 (58.8) | 23 (62.2) | 0.34 | 22 |
| - <200 | 39 (26.5) | 30 (29.4) | 9 (20) | 0.31 | 21 (27.6) | 10 (29.4) | 8 (21.6) | 0.72 | 22 |
| Haemoglobin (g/L) | 128.5 (114-144) | 130 (114-146.2) | 126 (114-144) | 0.72 | 129.5 (113.7-144) | 126 (116-149.5) | 131 (114-144) | 0.90 | 1 |
| Lymphocytes (x10^9^/L) | 1 (0.75-1.5) | 1.1 (0.8-1.55) | 1 (0.52-1.37) | 0.15 | 1.1 (0.7-1.6) | 1.1 (0.8-1.6) | 1 (0.8-1.6) | 0.034 | 4 |
| Creatinine (μmol/L) | 84.4 (65.4-111.4) | 82.2 (65.4-106.1) | 96.4 (65.4-152) | 0.039 | 77.8 (66.3-101.7) | 91.9 (72.5-109.6) | 97.2 (68-152.9) | 0.018 | 1 |
| Urea (mmol/L) | 13.9 (8.9-22.2) | 11.9 (8.66-19.6) | 18.6 (10.5-33.8) | 0.004 | 11 (8.6-17.8) | 16.4 (8.9-22.8) | 20 (11.4-40.3) | 0.001 | 3 |
| AST (µkat/L) | 0.68 (0.38-1.06) | 0.6 (0.43-0.91) | 0.82 (0.64-1.35) | 0.035 | 0.58 (0.42-1.07) | 0.83 (0.42-1.07) | 0.87 (0.6-1.39) | 0.005 | 1 |
| ALT (µkat/L) | 0.59 (0.35-0.89) | 0.57 (0.33-0.83) | 0.73 (0.45-0.97) | 0.077 | 0.57 (0.3-0.83) | 0.55 (0.40-0.85) | 0.75 (0.5-0.99) | 0.087 | 1 |
| GGT (µkat/L) | 1.14 (0.38-1.32) | 0.68 (0.33-1.25) | 0.83 (0.53-1.61) | 0.094 | 0.65 (0.32-1.17) | 0.92 (0.4-2.1) | 0.83 (0.5-1.25) | 0.21 | 23 |
| LDH (µkat/L) | 5.24 (3.91-7.95) | 5.04 (3.82-8.8) | 6.65 (4.33-8.8) | 0.006 | 0.83 (0.5-1.35) | 4.68 (3.64-6.93) | 5.63 (4.29-9.1) | 0.001 | 5 |
| Albumin (g/L) | 29.2 (24.5-33.37) | 29.2 (24.3-34.6) | 29.1 (25.9-31.1) | 0.63 | 30.7 (246-34.9) | 28.3 (24.3-34.6) | 27.2 (24.6-30.3) | 0.32 | 49 |
| Pro-BNP (ng/L) | 317 (5 (69.5-1573) | 193 (59-910) | 738 (193-2238) | 0.011 | 205 (55-733) | 232 (107-1788) | 826 (174-2306) | 0.035 | 45 |
| Troponin T (μg/L) | 15.7 (7.2-44.5) | 12.7 (6-42.6) | 19.5 (11.4-61.55) | 0.078 | 11.6 (5.4-35.3) | 20.6 (9.6-58.22) | 19.5 (9.9-67.45) | 0.038 | 36 |

AST aspartate transaminase; ALT alanine transaminase; LDH lactate dehydrogenase; pro-BNP precursor of the brain natriuretic peptide
